# Does altering antiretroviral therapy impact an individual’s gut microbiome?

**DOI:** 10.64898/2026.11.21.26368656

**Authors:** Gabriella Lindergard, Elizabeth Mann, Maximilian MacDonald, Andrew Ustianowski

## Abstract

**Introduction:** People with HIV are living with increased chronic inflammation with gut microbiome dysbiosis and microbial translocation emerging as contributing factors to inflammation and development of HIV associated co-morbidities. Single time-point cross-sectional population studies of individuals on antiretroviral drug therapy have suggested that some classes of antiretroviral therapy (ART) affect the gut microbiome more than others. There is, however, a lack of data examining the microbiome changes prospectively in individuals before and after alteration of their antiretroviral regiment. This study prospectively assesses changes in individual’s bacterial gut microbiome when switching from one antiretroviral drug or delivery method to another class or delivery method.

**Methods:** We conducted an observational prospective study of the gut microbiome in virologically controlled HIV infected individuals who were going through a planned switch of their ART regimen. We enrolled 15 participants and stool samples were collected before and 3-6 weeks after the switch of ART and analysed via shallow shotgun metagenomics.

**Results:** We did not detect a statistically significant overall difference in microbiome perturbation between the study participant group that switched their antiretroviral therapy compared to the controls who continued with the same ART during the timeframe of the study. Though underpowered to detect changes in subgroups, a noticeable observational shift in the 20 most abundant bacterial species was seen when individuals changed from a protease inhibitor-based regime to a non-nucleoside reverse transcriptase inhibitor (NNRTI) based regimen. Individuals who changed from NNRTI based treatment to integrase inhibitor-based treatment and from integrase inhibitor-based ART to protease inhibitor-based ART demonstrated numerical changes and shifts that failed to meet statistical significance. The initiation of intramuscular delivery of ART and a move away from oral drug did not result in a significant shift in the bacterial community in stool samples in this limited number of individuals.

**Conclusion:** We did not see statistically significant changes that were larger than those observed in controls in this small study. Switching from protease inhibitor to NNRTI demonstrated the largest shift in bacterial species that had been previously associated with positive health outcomes, indicating that protease inhibitor-based regimen might have a negative effect on the gut microbiome. Further larger prospective studies are planned to explore this area.

## Introduction

Gut health is increasingly recognised as being of importance for all aspects of human physical and mental wellbeing. In recent years there has been much interest and increased understanding of the importance of the gut microbiome and microbial translocation as factors in the development of HIV associated co-morbidities (Xiao et al. 2022; Bandera et al. 2018; Bai, Sonnerborg, and Nowak 2024; Cai et al. 2024; Pan, Wu, and Jin 2023; Islam et al. 2024; Mac Cann et al. 2024). Studies to date have largely been single time-point cross-sectional and we aimed to add to this collective understanding by prospectively investigating whether changes of the gut microbiome occur when individuals living with HIV change their antiretroviral treatment.

In 2024, 40.8 million people were living with HIV, of which 31.6 million were on antiretroviral therapy (ART) (https://www.unaids.org/en/resources/fact-sheet). The main purpose of ART is to suppress HIV viral replication to a level that is undetectable in peripheral blood and to thereby improve immune function. ART has been instrumental in changing the prognosis of HIV infection from a progressive lethal infection to a chronic and manageable infection. At this time there are seven distinct classes of antiretroviral drugs approved by the Food and Drug Administration (FDA); nucleoside reverse transcriptase inhibitors (NRTIs), non-nucleoside reverse transcriptase inhibitors (NNRTIs), protease inhibitors, integrase strand transfer inhibitors (INSTIs), chemokine receptor antagonists, fusion inhibitors, and now also a capsid inhibitor. These different antiretroviral (ARV) drug groups target different phases of the HIV replication cycle, from the entry into the cell, to reverse transcription, integration, and maturation stages.

The gut dysbiosis seen in untreated HIV infection is profound (Bandera et al. 2018; Pan, Wu, and Jin 2023). HIV affects the balance of normal host-microbe interactions both by changing the microbiome species and by disrupting normal host response mechanisms. However, this disrupted balance does not seem to be fully restored by the initiation of ART even though there is a move towards a microbiome more similar to that of HIV-uninfected individuals (Gaspar et al. 2024). This is observed even after long term

ART treatment with undetectable viral count where the bacterial make-up and abundance remain different from HIV negative controls possibly as a consequence of HIV reservoirs in, among other places, gut CD4^+^ cells (Ishizaka et al. 2024; Imahashi et al. 2021). Antiretroviral drugs could further contribute to this dysbiosis by directly altering the gut virome and this having indirect anti-bacterial and anti-bacteriophage effects with consequences for the individual (Villoslada-Blanco et al. 2022; Cao et al. 2022; Mukhopadhya et al. 2019; Scarpellini et al. 2015). An impaired intestinal barrier permits translocation of bacteria into the bloodstream and leads to a persistent low-grade background systemic inflammation. Despite new and effective oral ART drugs with better tolerability and safety, many individuals on ART suffer from gastrointestinal symptoms including diarrhoea, nausea and abdominal pain (Dikman et al. 2015; Clay and Crutchley 2014; Santos ASEAC 2019) and many complain of unexplained weight gain. Obesity has been associated with microbiome bacterial phylum-level changes, reduced bacterial diversity, and altered metabolic pathways and the obesity trait itself can be transmitted via microbiota transplantation (Turnbaugh, Hamady, et al. 2009; Turnbaugh, Ridaura, et al. 2009). There is also increasing evidence of the effect the microbiome has on host immune function, and dysfunction, and how its metabolites can affect and regulate cellular and molecular components such as macrophage metabolism (Zheng, Liwinski, and Elinav 2020; Rooks and Garrett 2016; Scott et al. 2022).

Recently, ART as an intramuscular injection once every one to two months has been approved in the UK and other nations (SOLAR study https://clinicaltrials.gov/ct2/show/study/NCT04542070). It is unknown what effect bypassing the gut in this way would have on the gut microbiome, but it is plausible that a lower drug concentration in the gut would decrease any negative impact on the gut microbiome.

A relatively low number of studies have examined the effect of ART on the gut microbiome. The vast majority of these have been population level analysis of single time-point cross sectional studies with multiple confounders and there have been very few examining the impact prospectively in individuals. In the available data, different combined ARTs have been associated with different profiles of gut microbiota (Villanueva-Millán et al. 2017; Villoslada-Blanco et al. 2022; Gaspar et al. 2024). When gut microbiota composition from these groups were analysed, primarily using 16S rDNA sequencing, investigators could detect different levels of bacterial diversity in the various groups, In short, current literature suggest that integrase strand transfer inhibitors are linked to better recovery and NNRTI based treatments are associated with proinflammatory bacteria. Functional studies, linking phylum changes of the microbiome with changes in immune function are still missing and we still need to understand the specific impact of each ART combination and whether the observed microbiota changes could indirectly contribute to immune reconstitution. It seems plausible that by improved understanding of the impact of changing ART we might be able to improve gut dysbiosis and help to better restore a healthy microbiota diversity and, as a result, improve the ongoing low-grade inflammation that contributes to HIV pathogenesis.

In this study we explore the effect that ART drugs have on an individuals’ microbiome by characterizing the intestinal microbiota in virologically suppressed individuals before switching from one drug class or mode of delivery to another. We compared this with samples collected from the same individual after 3-8 weeks on a stable regimen.

Comparing samples from each individual with themselves helps to eliminate the high variability seen between individuals resulting from lifestyle factors like diet, drugs, sexual habits, gender, and environment. The control group consisted of patients who did not switch ART but continued with their original medication.

## Materials and Methods

### Study design

We conducted an observational prospective study of the gut microbiome in virologically suppressed HIV infected individuals who were going through a planned switch of their ART regimen. We enrolled 15 participants over a period of 10 months, and individual participants were followed up once 3-6 weeks after the switch of ART had taken place.

Participants were identified through their usual care team as potentially eligible for the study and referred to the hospital research team who carried out eligibility checks and consent procedure. A basic medical history was conducted and information about current viral load, CD4^+^ count, time of HIV infection and time of treatment start, their normal diet and level of usual exercise, antibiotic use over the last 12 months and any current co-morbidities and concomitant medications. A serum blood sample was collected for future analysis, and patients were instructed to collect a stool sample and return it the next day. The time between blood sample and stool sample was never more than 24 hours. This first sample collection took place the same day before the first dose or the day before the ART switch.

### Study Participants

Patients were recruited into the study at North Manchester General Hospital, Manchester, UK, if they had been on stable antiretroviral treatment for at least 12 months, had an undetectable viral load (<50 copies per ml of blood) during the last 6 months and there was an imminent switch planned from one class of drugs or delivery method to another. 10 participants with a planned switch from one class of antiretroviral drugs to another class or delivery method were included and, in addition, 5 control participants who were HIV positive but did not change their ART regimen were also included in the study. Of the 15 study participants, two changed from oral cabotegravir and rilpivirine to injectable long-acting cabotegravir and rilpivirine (LA-CAB/RPV), three changed from NNRTI to Integrase inhibitor, two changed from Protease Inhibitor to NNRTI, one changed from Protease Inhibitor to Integrase Inhibitor, and one changed from Integrase Inhibitor to Protease Inhibitor. Of the controls, two were on stable Integrase Inhibitor, two on Protease Inhibitor and one on stable NNRTI with no change of their ART regimen.

### Assessments

All study assessments were collected at baseline and again around 4 weeks after the switch from one class of drugs or delivery method to another.

### Stool sample collection and processing

Study participants collected stool into 20ml sampling tubes and 2ml aliquots were frozen at -80°C within 8 hours of collection.

### DNA Extraction, PCR, Sequencing, and Sequence Processing

DNA extraction, library preparation, and bioinformatic processing were performed by Microbiome Insights (Vancouver, BC, Canada). DNA was extracted using the NucleoMag DNA Microbiome Kit (Macherey-Nagel, Düren, Germany) on a KingFisher automated extraction system. DNA concentration was quantified using a Qubit 4.0 fluorometer (Thermo Fisher Scientific, Waltham, MA, USA). This kit uses magnetic beads to specifically capture DNA while excluding organic inhibitors. In-house testing has shown that this kit has a good balance of DNA yield and quality, as demonstrated on a variety of environmental sample types.

Sequencing libraries were prepared using an Illumina Nextera library preparation kit following an in-house protocol (Illumina, San Diego, CA, USA). Libraries were sequenced on an Illumina NovaSeq X platform (300 cycles).

Raw sequence data were processed to remove adapter sequences, trim low-quality bases, and filter low complexity reads. Host-derived sequences were removed prior to taxonomic classification using Kraken2 with the PlusPF database. This cleaned bacterial prevalence data was supplied by Microbiome Insight Inc.

### Analysis

This was a pilot study aiming to identify changes in the diversity and abundance of the gut microbiome following a change in antiretroviral drug class or delivery method. Visualisation of the results was done with pandas (http://pandas.pydata.org) and matplotlib (https://matplotlib.org) using the cleaned bacterial prevalence data. The data frame underwent further cleaning and reordering for easier processing.

The 20 Most Abundant bacteria were chosen by selecting the 20 bacteria with the highest average prevalence across the cleaned data frame from the whole cohort. Largest relative difference was calculated using log2 fold-change to equally weight negative change with positive change, where a difference of +1 represents a 2-fold increase and -1 a 2-fold decrease. Prior and post values of 0 are excluded from this visualisation as they are not compatible with the metric.

We also visualised changes to a panel of bacterial species identified to be showing the strongest overall correlation with a selection of markers of nutritional and cardiometabolic health (Asnicar et al. 2021)

### Ethical Considerations

The study received a favourable ethical opinion from the Greater Manchester South Research Ethics Committee (REC reference 24/NW/0009, IRAS project ID 333914) and followed the principals set out in the UK Policy Framework for Health and Social Care Research.

## Results

The overall number of bacterial species identified and classified for each of the collected stool samples were around 9300. We chose to analyse the changes of the 20 most abundant bacterial species for each of the samples and, in addition, to specifically describe changes to a panel of bacterial species identified to be showing the strongest overall correlation with a selection of markers of nutritional and cardiometabolic health (Asnicar et al. 2021)

Due to the small sample size of this pilot study, the impact of individuals changing their ARV drug treatment did not present statistically significant changes in their identifiable bacterial gut microbiome compared to individuals who were on a stable antiretroviral therapy regardless of the class of drugs or delivery method (Figure 1 and Figure 2) within the timeframe of the study. Despite not showing statistical significance there are several interesting trends.

**Figure 1.**
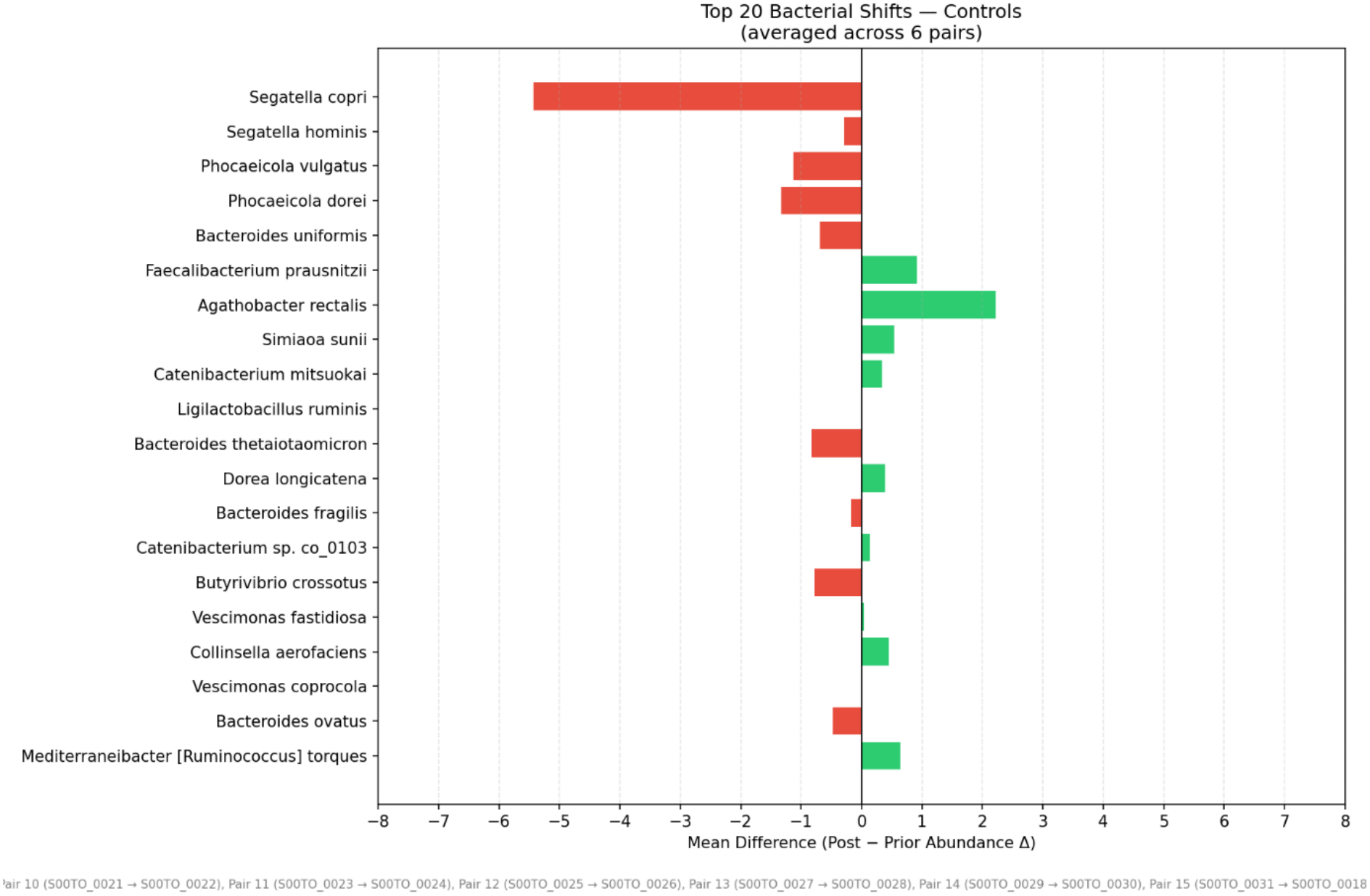
Increase (green) and decrease (red) of the 20 most abundant bacterial species in stool samples from control participants who had not changed treatment.

**Figure 2.**
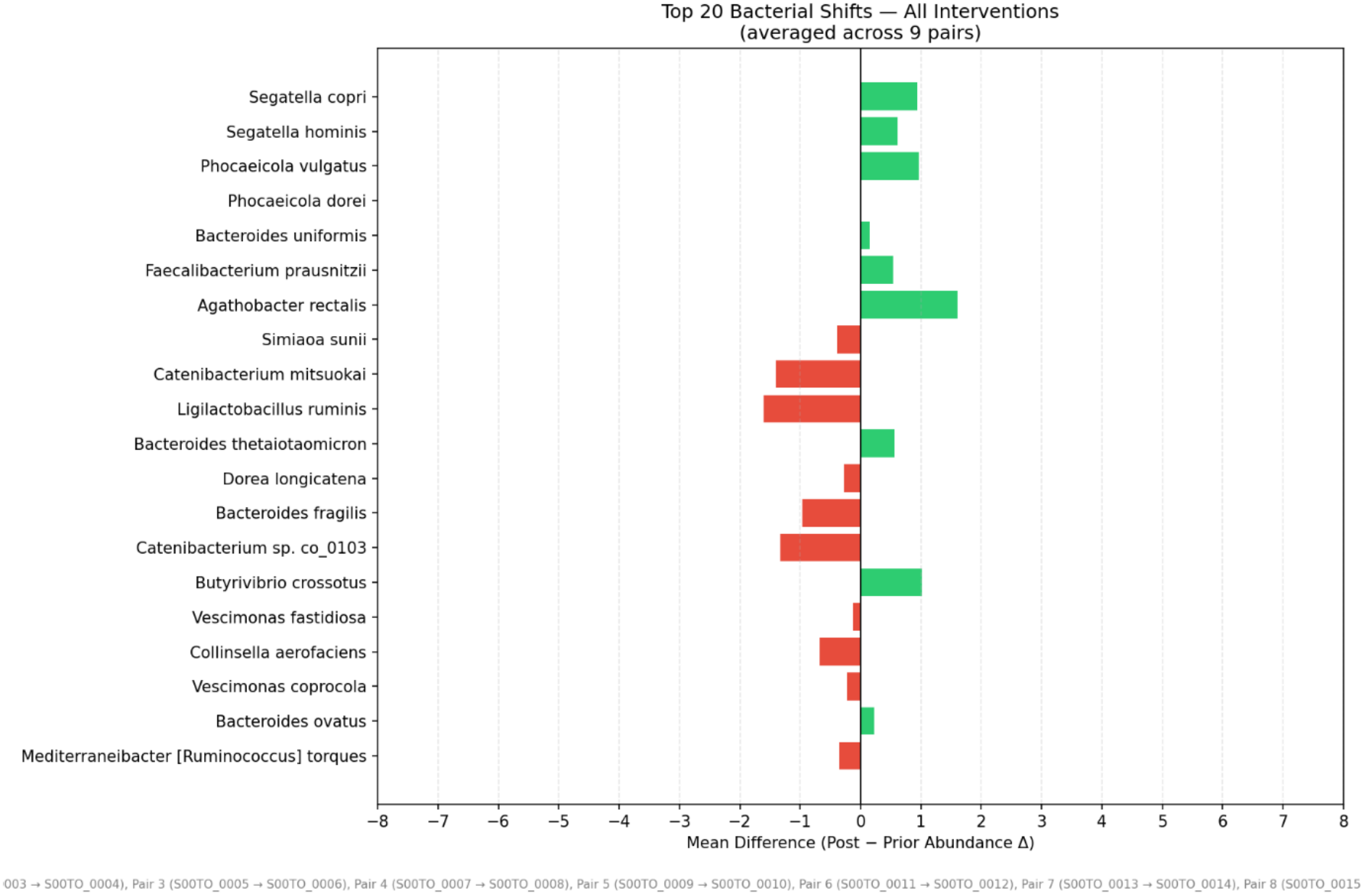
Increase (green) and decrease (red) of the 20 most abundant bacterial species in stool samples from study participants who changed the class or delivery method of their ART.

The biggest shifts seem to be when changing from a protease inhibitor-based regime to a NNRTI based regiment (Figure 3) whereas the least change seem to be after a switch from protease inhibitor ART to integrase inhibitor ART (Figure 4). A change from NNRTI based treatment to Integrase inhibitor-based treatment and Integrase inhibitor-based ART to protease inhibitor-based ART resulted in an intermediate change in the 20 most abundant bacterial species in the stool samples. It was noted that the initiation of intramuscular delivery of ART and a move away from oral drug did not result in a significant shift in the bacterial community in the stool samples.

**Figure 3.**
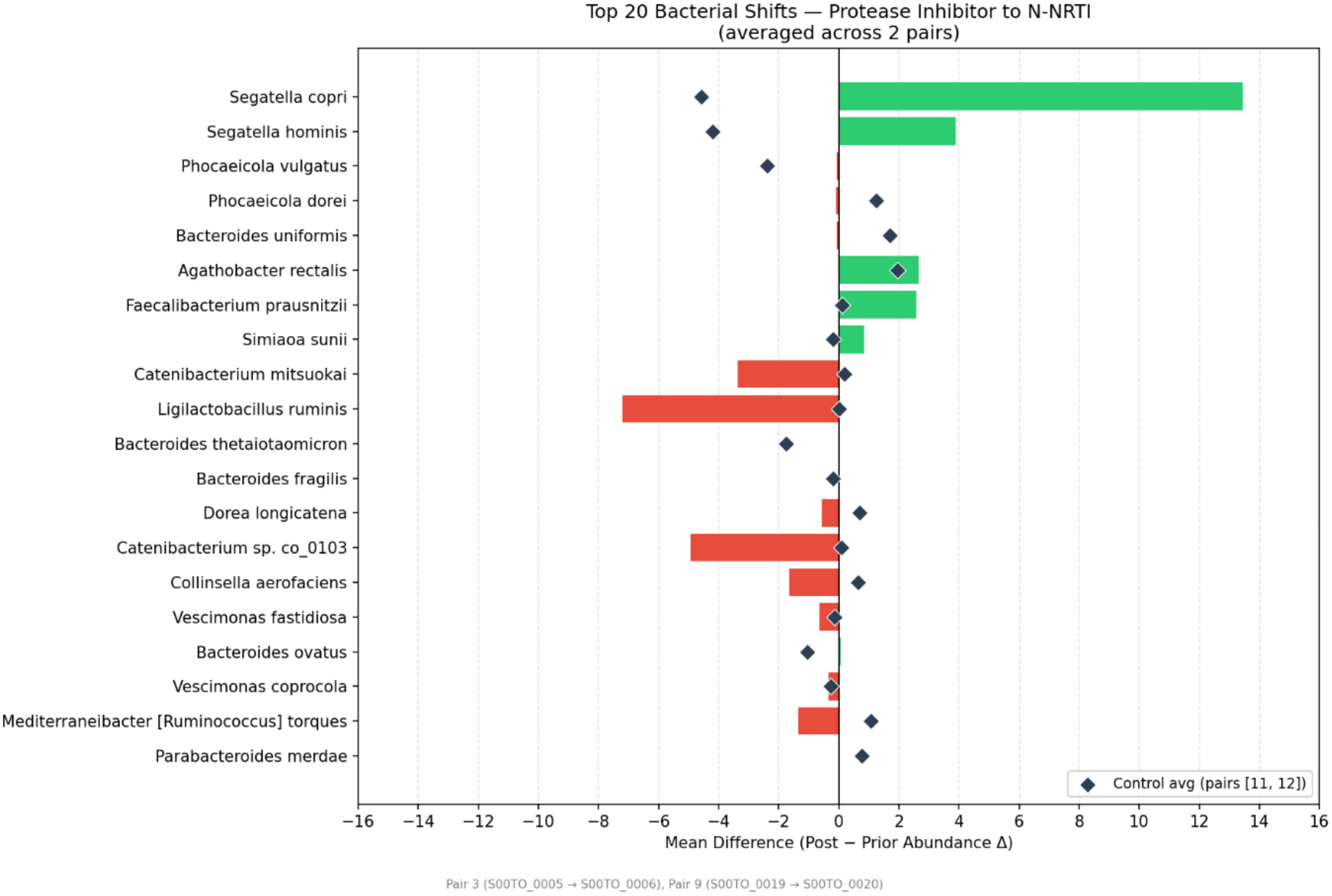
Increase (green) and decrease (red) of the 20 most abundant bacterial species in stool samples from study participants who changed from a protease inhibitor-based regime to a NNRTI based regiment.

**Figure 4.**
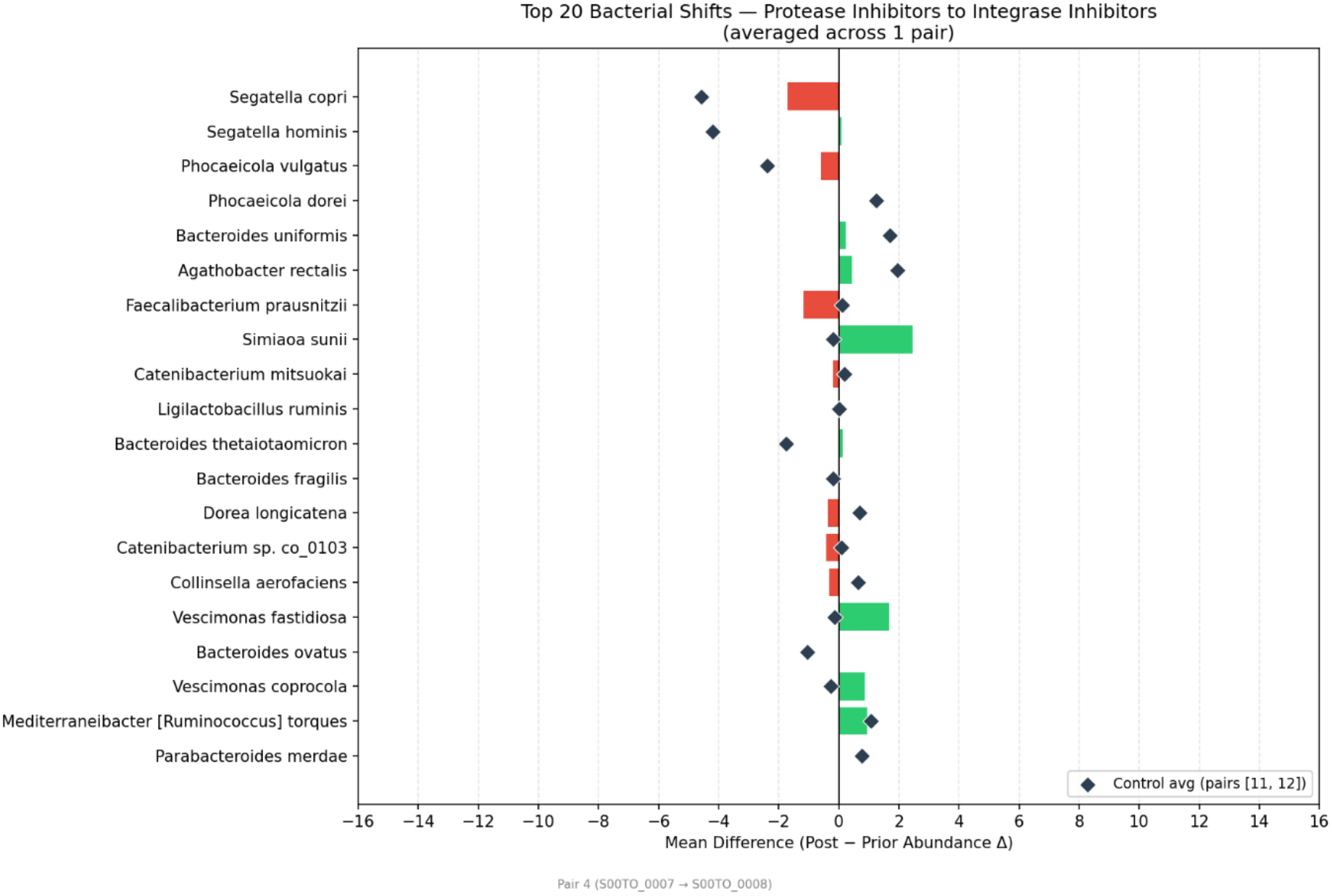
Increase (green) and decrease (red) of the 20 most abundant bacterial species in stool samples from study participants who changed from protease inhibitor ART to integrase inhibitor ART.

Analysis of the results from the PREDICT 1 study (Asnicar et al 2021) lead to a description of microbial signatures of cardiometabolic health and the identification of bacterial species associated with positive and negative indicators of cardiometabolic health. Of the 15 positively and 15 negatively associated bacterial species listed in the paper, our sequencing analysis method was able to identify 7 of the positive and 9 of the negative bacterial species. The largest shift towards the positively linked bacterial species were seen after a change from protease inhibitors to N/NRTIs (Figure 5) where there was an increase in the beneficial bacteria *Segatella copri, Faecalibacterium prausnitzii* and *Haemophilus parainfluenzae* as well as a decrease in the negatively associated *Escherichia coli*. The opposite was seen after a change from integrase inhibitor to protease inhibitors which resulted in a decrease in *Segatella copri* and *Faecalibacterium prausnitzii*, both regarded as beneficial bacteria. The change from protease inhibitor to integrase inhibitor did, however, not display obvious alterations and the changes seen when changing from N/NRTI to integrase inhibitor was very small even though the changes seen in all 3 of the bacterial species were in a positive direction.

**Figure 5.**
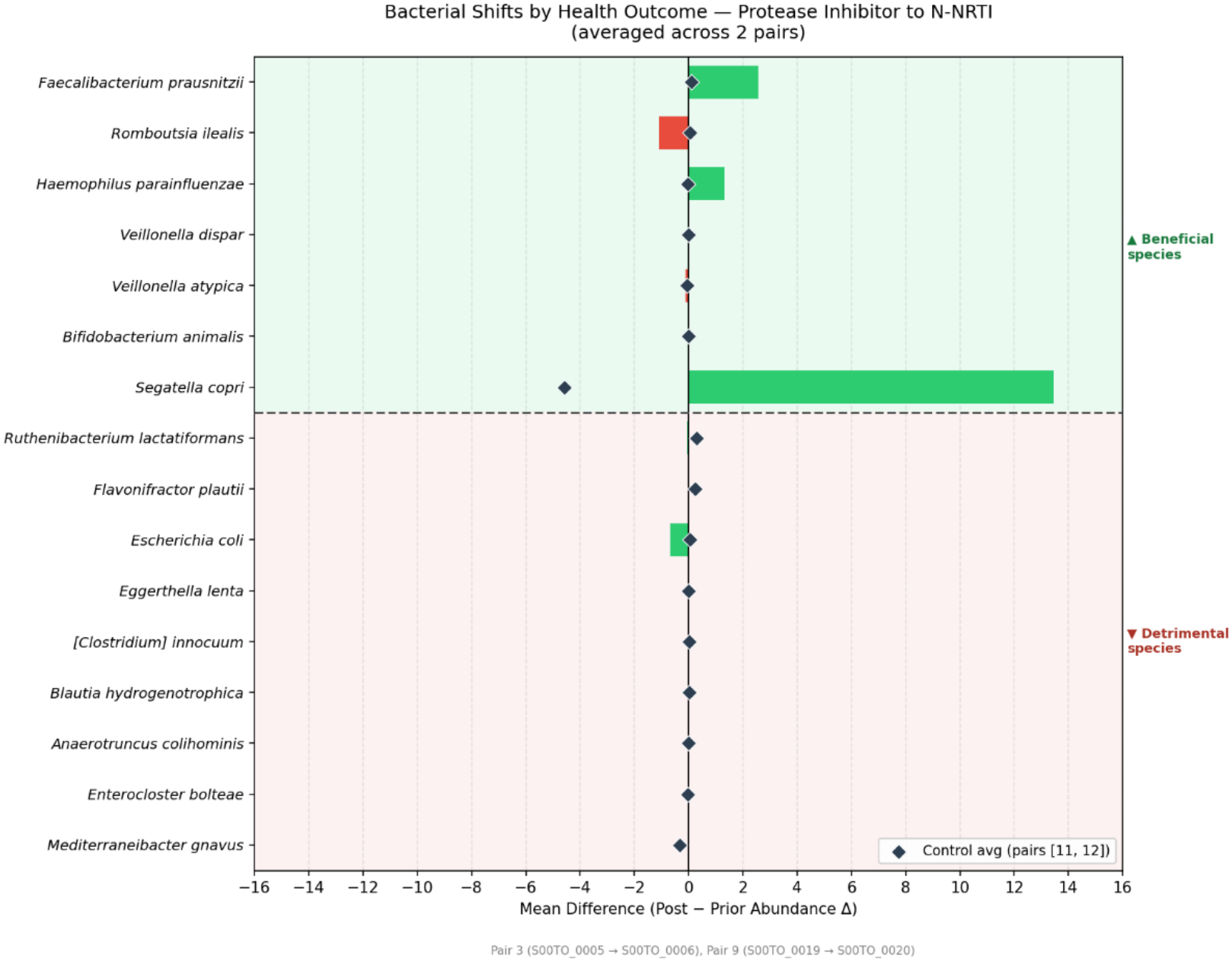
Increase (green) and decrease (red) of 7 bacterial species associated with positive cardiometabolic health outcomes and 9 bacterial species associated with negative cardiometabolic health outcomes in samples from study participants changing from protease inhibitors to N/NRTIs.

The analysis was also extended to include relative change within all the identified species, but no pattern or consistent changes could be noted within the small subgroup. One interesting result could be seen in a sample from an individual changing their ART from NNRTI based to integrase inhibitor based resulting in a marked decrease of *Akkermansia muciniphila*, which is often cited as a beneficial bacterium and much promoted for commercial interest (Figure 6).

**Figure 6.**
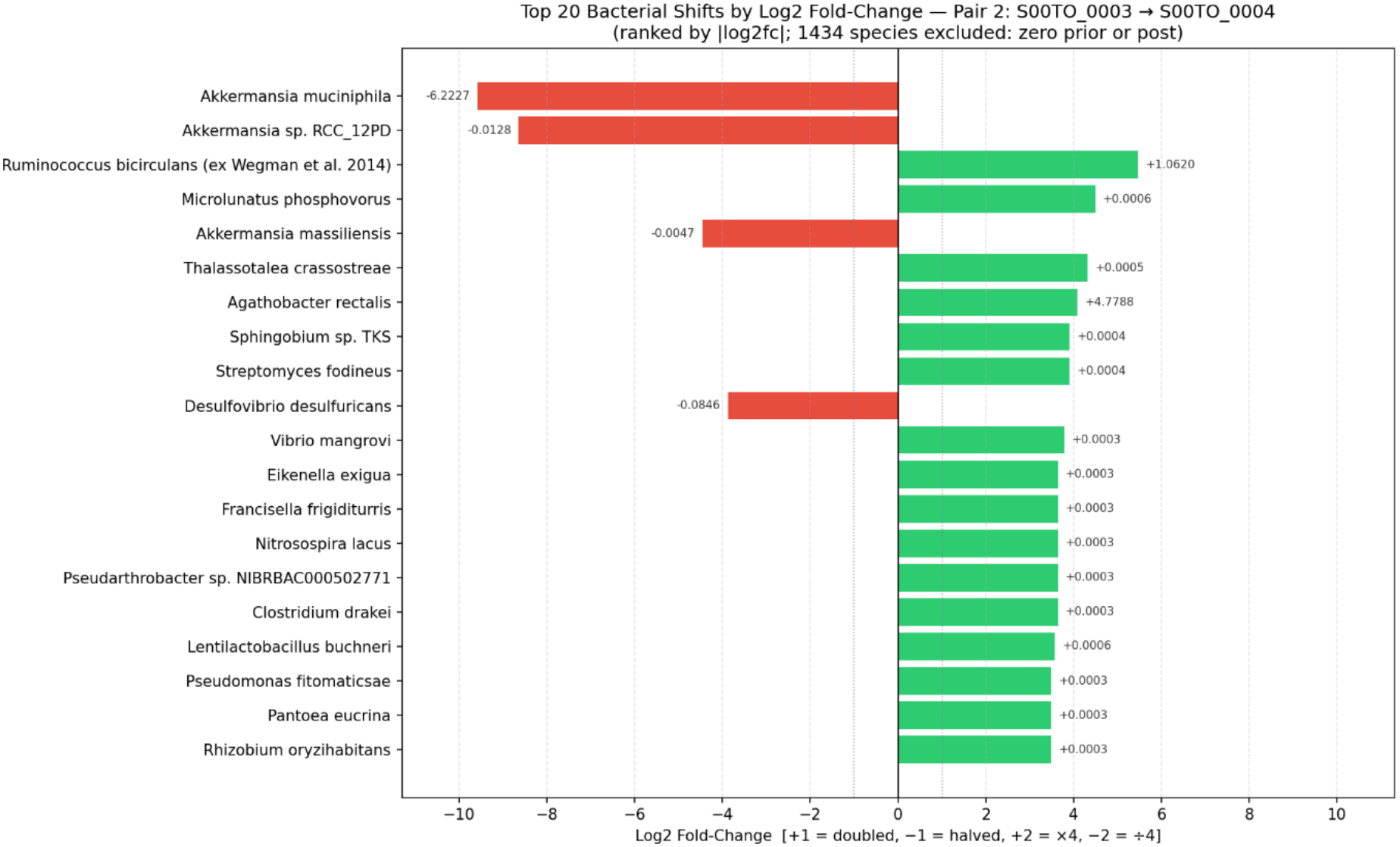
Relative change in sample from an individual changing their ART from N/NRTI-based regimen to integrase inhibitor-based regimen.

## Discussion

Much of the available gut microbiome sequencing data in the literature to date is the result of 16S amplicon sequencing. But as we are seeing a transition towards the use of shotgun metagenomics, we instead chose to use a shallow shotgun sequencing approach to benefit from the enhanced taxonomic classification. This resulted in a fairly high percentage of unidentified bacterial species, mostly around 3% but in some samples reaching up to 20%. During the analysis, we choose not to normalize the data as we were still interested in identifying change rather than total abundance. Because of the small sample size, the more commonly used Shannon index to compute alpha diversity was less suitable for describing the richness and evenness of the gut microbiome and we have visualised the data in the shape of diverging bar charts instead. The aim of this pilot study was to identify areas of interest for further studies as it has not yet been established if all classes of antiretroviral drugs have the same effect on the gut microbiome. We did not see clear significant changes that were larger than that seen in the control recruits in our study but there were nevertheless some potentially interesting findings that could be used as guides for future studies. The change from protease inhibitor to NNRTI had the largest shift in bacterial species towards more beneficial bacterial species, indicating that protease inhibitor-based regimen might have a negative effect on the gut microbiome. A follow-on study could concentrate on this patient group and recruit a larger number of participants. It would also be valuable to include a longer time frame and sample at further intervals to better capture natural fluctuations in the abundance of specific bacterial species present in the gut.

## Data Availability

All data produced in the present study are available upon reasonable request to the authors.

## Acknowledgement

We are very grateful to all the individuals who chose to participate in this study and gave their time to further our understanding on what effect antiretroviral drugs have on the gut microbiome. We would also like to thank the Infectious Diseases Research Team at North Manchester General Hospital and Dr Thomas Lamb for help with recruitment.

The study was funded by a University of Manchester Clinical Lecturer Grant.

